# An AI-Powered tissue-agnostic cellular morphometrics biomarker for risk assessment in patients with pan-gastrointestinal precancerous lesions and cancers

**DOI:** 10.1101/2024.11.14.24317353

**Authors:** Pin Wang, Chengfei Jiang, April W. Mao, Qi Sun, Hong Zhu, Jamie Inman, Susan Celniker, Antoine M. Snijders, David W Threadgill, Allan Balmain, Bo Hang, Jia Fan, Jian-Hua Mao, Lei Wang, Hang Chang

**Affiliations:** Department of Gastroenterology, Nanjing Drum Tower Hospital, Affiliated Hospital of Medical School, Nanjing University, Nanjing, Jiangsu, 210000, China; Department of Liver Surgery and Transplantation and Key Laboratory of Carcinogenesis and Cancer Invasion, Ministry of Education, Liver Cancer Institute, Zhongshan Hospital, Fudan University, Shanghai 200032, China; Department of Mathematics, University of California Los Angeles, Los Angeles, CA, 90095, USA; Biological Systems and Engineering Division, Lawrence Berkeley National Laboratory, Berkeley, CA, 94720, USA; Department of Pathology, Affiliated Drum Tower Hospital, Medical School of Nanjing University, Nanjing, China; Department of Gastroenterology, The First Affiliated Hospital of Nanjing Medical University, Nanjing 210029, China; Berkeley Biomedical Data Science Center, Lawrence Berkeley National Laboratory, Berkeley, CA, 94720, USA; Department of Nutrition, Texas A&M University, College Station, TX, 77843, USA; Department of Molecular and Cellular Medicine and Department of Biochemistry & Biophysics, Texas A&M University, College Station, TX, 77843, USA; Helen Diller Family Comprehensive Cancer Center, University of California San Francisco, San Francisco, CA94158, USA; Department of Biochemistry and Biophysics, University of California San Francisco, San Francisco, CA 94518, USA; Institutes of Biomedical Sciences, Fudan University, Shanghai 200032, China; State Key Laboratory of Genetic Engineering, Fudan University, Shanghai 200433, China

**Keywords:** Tissue-agnostic biomarker, Gastrointestinal cancer, Precancerous lesions, Artificial intelligence (AI), Cellular morphometrics biomarker (CMB), whole-slide images (WSI), CMB risk score (CMBRS), CMB risk group (CMBRG), Multimodal integration

## Abstract

**PURPOSE:** Tissue-agnostic biomarkers that capture the commonality in cancer biology, may provide a new avenue for treatment development and optimization across cancer types. Here, we aimed to evaluate and validate the clinical value of a tissue-agnostic cellular morphometrics biomarker (CMB) signature, which was discovered by artificial intelligence (AI) from H&E-stained whole-slide images (WSI) of diagnostic slides of colon cancers, in pan-gastrointestinal (pan-GI) pre-cancer lesions and cancers.

**METHODS:** We discovered CMBs from WSI using our well-established CMB-ML pipeline and established a CMB risk score (CMBRS) using multivariate regression models. Based on CMBRS, we assigned individual patients from The Cancer Genome Atlas Colon Adenocarcinoma Cohort (TCGA-COAD) (n=430) to CMB risk groups (CMBRG). We then extensively evaluated tissue-agnostic clinical value of CMB signature, CMBRS and CMBRG in multi-cohorts with different types of GI cancer (n=2,219) and risk assessment of precancerous lesions (n=1,016). We unraveled each CMB-related biological function using bulk RNA-sequencing, single-cell RNA-sequencing (scRNA-seq) and opal multiplex immunohistochemistry (IHC) techniques.

**RESULTS:** From the TCGA-COAD cohort, we developed a 13-CMB signature and constructed CMBRS/CMBRG that predict prognosis of colon cancer patients. Importantly, this 13-CMB signature proved prognostic and predictive values for TCGA patients with rectal, gastric and esophageal cancer independent of traditional clinical factors. These findings were independently validated using multiple cohorts from Drum Tower Hospital. Moreover, 13-CMB signature exhibited the power for risk stratification of colon adenoma and early esophageal neoplastic lesion patients for predicting cancer progression. In addition, we demonstrated and validated independent prognostic impacts of gene signatures and CMB signatures and a significant increase in predictive power by integration of CMB signature, gene signature and clinical factors. Correlations between CMBs and gene expression levels revealed the association of each CMB with biological functions including cell proliferation, epithelial-to-mesenchymal transition and immune microenvironment. The association of CMBs with the immune microenvironment was prospectively validated by scRNA-seq and was further confirmed by Opal multiplex IHC staining in colon cancer.

**CONCLUSION:** This study demonstrates the clinical value of tissue-agnostic AI-empowered CMB signature from WSI with defined biological functions, which can be used in clinical settings to assess risk, diagnose disease, and guide clinical interventions. Tissue-agnostic CMBs potentially provide a new avenue for a rapid, robust and cost-effective cross-cancer prediction that is essential for developing common treatment strategy for multiple cancers.

## Introduction

The past decades of cancer research and clinical oncology have been devoted to tumor characterization at both the cellular and molecular levels, which led to the emerging evidences of tissue-agnostic treatment benefits of certain drugs (e.g., nivolumab, a PD1 inhibitor ^1^, trastuzumab deruxtecan, a HER2 targeted antibody ^2,3^ and Olaparib, an polymerase inhibitor with activity in germline BRCA1 and BRCA2 ^4^), leveraging the molecular profiling of tumors regardless of the organ in which tumors are originated. Moreover, most cancer types can be further subdivided into different molecular subgroups, including breast cancer ^5^, lung cancer ^6^, brain cancer ^7^ and gastric cancer ^8,9^, that have distinct clinical outcomes and requires subtype-specific treatment optimization. In sight of these evidences, regulatory agencies, including the US Food and Drug Administration (FDA), are working towards the guideline on tissue-agnostic drug development ^10,11^, with the potential to reclassify cancer and restructure oncology based on their molecular landscape.

However, the intertumoral and intratumoral cancer heterogeneity is not solely defined by their molecular profile, but also reflected in tissue histology that captures dynamic microenvironments including the cytoplasm, nucleus, organelles and extra-cellular components, which therefore remains as the gold standard for cancer diagnosis and together contribute to diverse therapeutic responses ^12,13^. In addition, the affordability, accessibility and the turnaround time of genomic profiling are practical barriers to the implementation of genomics as a tool in primary care, which not only impedes its global application ^14,15^ but also exacerbates health disparity ^15^. In contrast, histopathological slides with hematoxylin and eosin (H&E) staining are routinely used in cancer diagnosis by pathologists. Therefore, the tissue-agnostic characterization and biomarker development of tumors at pathologic level is believed to provide a new avenue to overcome the challenges in precision oncology.

With the adoption of digital workflows in histopathology and recent advancements in artificial intelligence (AI), numerous studies have revealed the possibility to discover cancer biomarkers in the form of whole slide images (WSI)^16^. And we previously developed and extensively validated a powerful AI-based pipeline: Cellular Morphometric Biomarker (CMB) via Machine Learning (CMB-ML), to profile the cellular morphometric landscape from WSI in multiple type of cancers and model systems ^17–19^ that have demonstrated association with prognosis, treatment response and tumor microenvironments. However, the majority effort in tissue-histology-based biomarker development is still devoted into specific cancer type, leaving the potential of tissue-agnostic biomarker insufficiently explored, which is essential to the tissue-agnostic cancer diagnosis, treatment and drug development.

Gastrointestinal (GI) cancers account for a quarter of the global cancer incidence and a third of cancer-related deaths, and the disparities across countries warrants context-specific targeted GI cancer control and health systems planning ^20^. Tremendous efforts have been devoted to the molecular profiling and biomarker development both cancer-type-specifically and tissue-agnostically in GI cancers ^21–24^. However, the progress in the management and treatment of GI precancers and cancers has been limited ^25^, which imposes significant clinical challenges and underscores the unmet needs in precision oncology. In this study, we conducted comprehensive evaluation and validation of clinical value of an AI-empowered tissue-agnostic CMB signature discovered from WSIs of colon cancers in pan-GI pre-cancers and cancers (Figure 1). Specifically, we first discovered CMBs from WSI and established a CMB risk score (CMBRS) system and CMB risk group (CMBRG) in TCGA-COAD cohort and then extensively evaluated the clinical utility of CMB signature, CMBRS and CMBRG for GI precancerous patient risk assessment and GI cancer patient prognosis. Furthermore, the CMB-associated biological functions were assessed by bulk RNA-sequencing and validated by single-cell RNA-sequencing (scRNA-seq) and opal multiplex immunohistochemistry (IHC) techniques. Our findings demonstrate that tissue-agnostic CMBs can provide a new avenue for a rapid, robust and cost-effective aid to make clinical decisions in GI pre-cancer and cancer patients.

**Figure 1.**
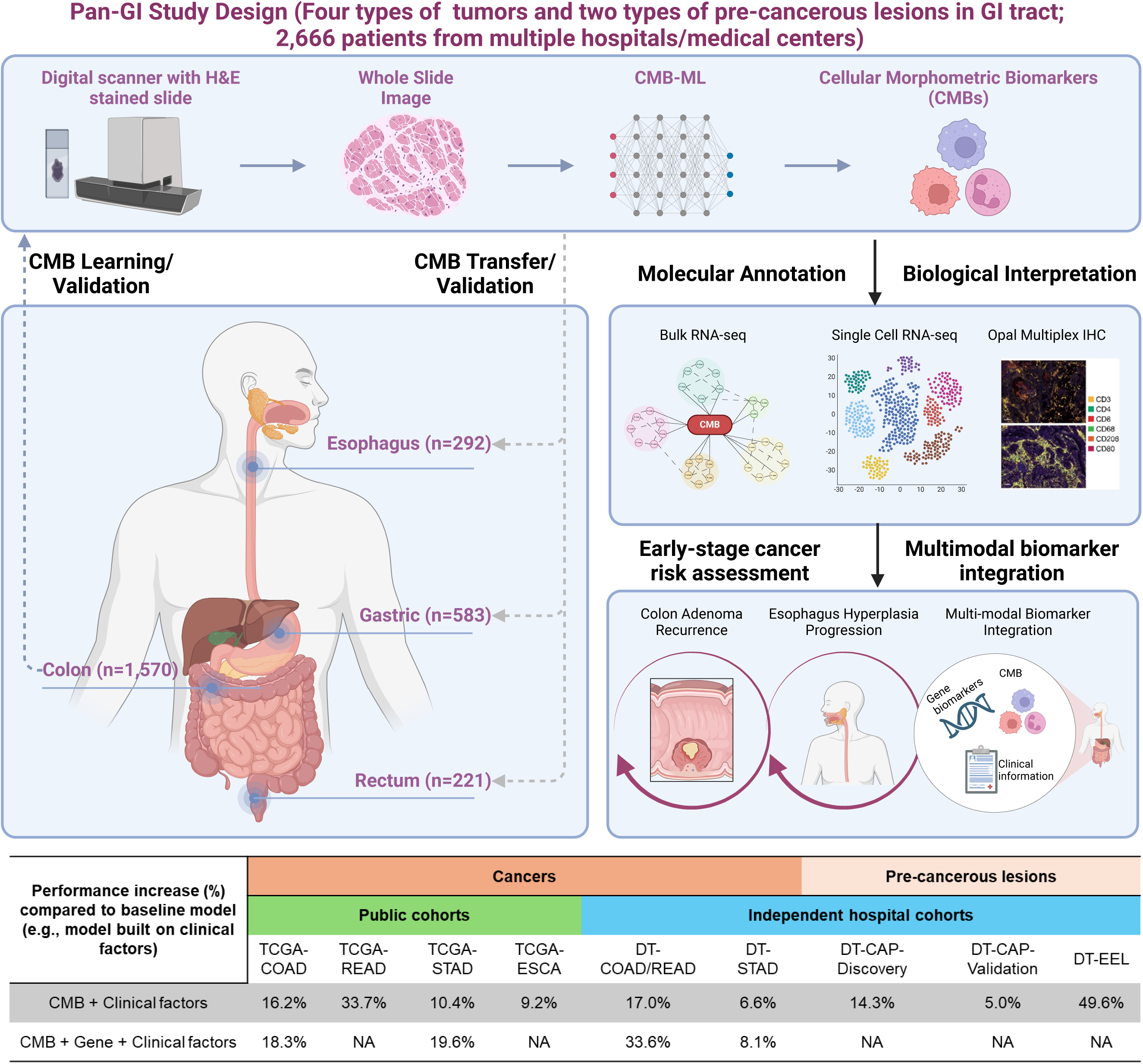
Pan-GI Study Design and Model Performance.

## Methods

### Human cohorts

The human gastrointestinal cancer and precancerous cohorts included in this study consisting of 1,118 patients from the Cancer Genome Atlas (TCGA) and 1,558 patients collected from the Nanjing Drum Tower Hospital (DT), the Affiliated Hospital of Nanjing University Medical School. Specifically, the TCGA-COAD cohort (Supplementary Table S1) included 430 patients diagnosed with colon adenocarcinoma, the TCGA-STAD cohort (Supplementary Table S2) included 375 patients diagnosed with stomach adenocarcinoma, the TCGA-READ cohort (Supplementary Table S3) included 157 patients diagnosed with rectal adenocarcinoma, and the TCGA-ESCA cohort (Supplementary Table S4) included 156 patients diagnosed with esophageal carcinoma. The inclusion criteria in TCGA cohorts are primary tumors with diagnostic slides and follow-up information.

The DT-COAD/READ cohort (Supplementary Figure S1, Supplementary Table S5 and S6) included 243 patients diagnosed with primary colon adenocarcinoma and 64 patients diagnosed with primary rectal adenocarcinoma, who received radical surgery between Jan. 2007∼Dec.2015 at Drum Tower Hospital. There were 177 (57.7%) male and130 (42.3%) female patients, with a median age of 60.0 years (range: 15-87 years). The complete cohort consisted of 1,135 patients, with 827 excluded due to loss of follow-up (n=542), lack of clinical information (n=152), diagnosis of other tumor between the follow-up duration (n=16), history of colorectal cancer before this study (n=21), no pathological slides or with insufficient quality slides (n=97).

The DT-STAD cohort (Supplementary Figure S2, Supplementary Table S7) included 208 patients diagnosed with primary gastric cancer, who received radical gastrectomy between Jan. 2009∼Jun.2015 at Drum Tower Hospital. There were 157 (75.5%) male and 51 (24.5%) female patients, with a median age of 62 years (range: 24-90 years). The complete cohort consisted of 1,198 patients, with 990 excluded due to loss of follow-up (n=766), lack of clinical information (n=67), history of gastric cancer before this study (n=77), no pathological slides or with insufficient quality slides (n=80).

The DT-CAP cohort (Supplementary Figure S3, Supplementary Tables S8-S10) included 880 patients received colonoscopy between Jan. 2013∼Dec. 2021 at Drum Tower Hospital. There were 565 (64.2%) male and 315 (35.8%) female patients, with a median age of 60 years (range: 21-91 years). The complete cohort consisted of 8831 patients, with 7951 excluded due to non-adenoma disease (n=3,896), lost follow-up (n=3,692), lack of clinical information (n=135), history of colectomy (n=30), with hyperplastic polyps (n=116), no pathological slides or with insufficient quality slides (n=82). The cohort was divided into two sets, DT-CAP-Discovery set (n=299) whose patients were diagnosed from Jan. 2013 to Dec. 2016 and DT-CAP-Validation set (n=581) whose patients were diagnosed from Jan. 2017 to Dec. 2021.

The DT-EEL cohort (Supplementary Figure S4, Supplementary Table S11) included 136 patients with early neoplastic lesions diagnosed by gastroscopy between Jan. 2013∼Dec. 2021 at Drum Tower Hospital. There were 86 (66.2%) male and 44 (33.8%) female patients, with a median age of 63 years (range: 34-85 years). The complete cohort consisted of 704 patients, with 568 excluded due to lost follow-up (n=237), the interval between two inspections less than 1 year (n=297), diagnosis of other tumors within the follow-up duration (n=8), history of esophagectomy (n=3), no pathological slides or with insufficient quality slides (n=23).

The DT-COAD-scRNASeq cohort (Supplementary Figure S5, Supplementary Table S12) included 20 patients with colon cancer and received radical surgery from Aug. 2023 to Oct. 2023. There were 10 (50.0%) male and 10 (50.0%) female patients, with a median age of 63.5 years (range:32-91 years). The complete cohort consisted of 17 patients, with 3 excluded due to insufficient sample quality (n=3). We analyzed gene expression, diagnostic slides and clinical data of human gastrointestinal cancer and precancerous lesions from two cohorts. The public cohort consists of 1,118 patients collected from TCGA database, and the validation cohort consists of 1,558 patients collected from the Nanjing Drum Tower Hospital, the Affiliated Hospital of Nanjing University Medical School. The validation study was approved by the Institutional Review Board of Nanjing Drum Tower Hospital and conducted in accordance with the Declaration of Helsinki. The model development using the publicly available TCGA-COAD cohort was performed at Lawrence Berkeley National Laboratory. The hospital validation study was approved by the Institutional Review Board (IRB) at the participating hospital and was independently carried out at Nanjing Drum Tower Hospital.

### Identification of CMBs and construction of the CMB Risk Score (CMBRS) and CMB Risk Group (CMBRG) from TCGA-COAD

Patients with H&E-stained diagnostic slides and complete clinical information were used to develop the CMBs, CMBRS and CMBRG. Based on the stacked predictive sparse decomposition (SPSD) ^26^ technique and our Cellular Morphometric Biomarker via Machine Learning (CMB-ML) pipeline ^17, 27^, we defined 128 CMBs from cellular objects extracted from the whole slide images (WSI) of H&E stained tissue histology sections in TCGA-COAD cohort. In the CMB-ML pipeline, we used a single network layer with 128 CMBs and a sparsity constraint of 30 at a fixed random sampling rate of 1000 cellular objects per WSIs from the cohort. The pre-trained CMB-ML model reconstructed each cellular region as a sparse combination of pre-defined 128 CMBs and thereafter represents each patient as an aggregation of all delineated cellular objects belonging to the same patient. The experimental settings was identical to our previous study^27^ to keep the reconstruction error less than 10% during training.

The prognostic effect of high or low levels of each CMB on overall survival (OS) was assessed by Kaplan-Meier analysis (survminer package in R, version 0.4.8) and log-rank test (survival package in R, version 3.2-3), where TCGA-COAD cohort was divided into two groups (i.e., CMB-high and CMB-low groups) based on each CMB (cut-off estimated using survminer package in R, version 0.4.8). The set of CMBs as a prognostic signature was selected via a multivariate Cox proportional hazards (CoxPH) regression model, including these CMBs with a significant effect on poor outcome events.

The construction of CMBRS was defined below, where the coefficients of the final CMBs as categorical variables were obtained from multivariate CoxPH regression analysis:

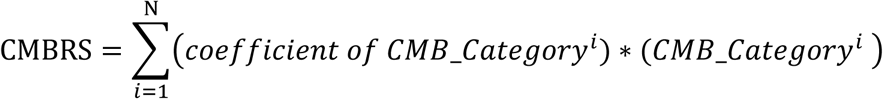

Where N is the number of final CMBs that were independently and significantly associated with poor outcome events, and *CMB*_*Category*^*i*^ is the category of the *i*^*t*ℎ^ CMB (i.e., CMB-high=1; CMB-low=0). After CMBRS construction, the retrospective cohort was divided into high-risk group (top 1/3), intermediate-risk group (middle 1/3), and low-risk group (bottom 1/3) based on CMBRS, and the CMBRS cut-offs between high/intermediate and intermediate/low were recorded and fixed for as the CMBRG model. During hospital validation study, both the cut-points for CMB and CMBRS remain unchanged.

### Re-optimization of CMBs

During biomarker transfer to different tumor types, the original CMBs developed from TCGA-COAD remain fixed. The cut-points of each CMB were re-optimized using the same strategy as for TCGA-COAD cohort, and the cut-point for CMBRS was then established accordingly. After re-optimization per TCGA cohort, these cut-points remain unchanged during hospital validation for the same tumor type.

### Functional annotation of CMB-correlated genes

For each CMB, Pearson correlation analysis was performed among all the TCGA samples described above to determine the genes significantly associated with each CMB (Statistical significance: *P* value < 0.05). The enrichment analysis was conducted using online database: Gene Ontology (GO) and Kyoto Encyclopedia of Genes and Genomes (KEGG) to annotate the biological function of the genes related to different CMB. (Statistical significance: adjusted p value < 0.05, cell component enrichment: *Q* value < 0.05, biological process enrichment *Q* value <0.2, molecular function enrichment: *Q* value <0.2, KEGG enrichment *Q* value < 0.2) The result of these enrichment analysis was applied to draw the network diagram of the same function shared with different CMB. The functional networks were then created in Cytoscape (version 3.10.1) based on the results of enrichment analysis for Cellular Components, Molecular Function, Biological Process and KEGG, respectively.

### Preparation of single-cell suspensions and scRNA sequencing

In this study we prospectively collected 17 colon cancer samples to validate our findings using scRNA sequencing, where all the patients have been diagnosed with colon cancer through biopsy before the resection. For isolation of colon cells, fresh intestines were collected from colorectal resection. Three biological replicates were included in each group. Intestines were dissected and washed to remove fecal content with pre-cold PBS for several times, minced into small pieces, and incubated with 0.5 mg/mL digestive enzymes 1 for 30 min at 37℃ in a water bath. After centrifugation, supernatant was collected and the precipitation was continued to be digestion with 0.5 mg/ml digestive enzyme 1 for another 30 min. After digestion for total 60 min, all the supernatant and precipitation were combined and filtered through a 70 μM cell strainer (BD Falcon, 352350). After centrifugation at 500 g for 5 min, the precipitation was further digested with 1 mL trypsin for 15 min. The supernatants were collected, then all the supernatant was filtered through 35 μM cell strainer (BD Falcon, 352235). After centrifugation at 500 g for 5 min, the cell precipitated was resuspension with PBS+0.01%BSA.

Single cell suspensions were loaded on 10x Genomics Chromium™ according to manufacturer’s protocol based on the 10x GEM Code proprietary technology. Single-cell RNA-Seq libraries were prepared using 10x Genomics Chromium Next GEM Single Cell 3ʹ Kits v3.1 according to manufacturer’s protocol. Briefly, the initial step involves performing an emulsion where individual cells were isolated into droplets together with gel beads coated with unique primers bearing 10x Genomics cell barcodes, unique molecular identifiers (UMI), and poly(dT) sequences. Reverse transcription reactions were engaged to generate barcoded full-length cDNA followed by the disruption of emulsions using the recovery agent and cDNA clean up. Bulk cDNA was amplified and cleaned up. Sequencing libraries were constructed using the reagents from the 10x Genomics Chromium Next GEM Single Cell 3ʹ Kits v3.1, following these steps: (1) fragmentation, end repair, and a-tailing; (2) size selection with SPRI select; (3) adaptor ligation; (4) post ligation cleanup with SPRI select; (5) sample index PCR and cleanup with SPRI select beads. Indexed libraries were pooled according to number of cells and sequenced on a NovaSeq 6000 (Illumina) using paired-end 150 bp.

### scRNA-seq data processing

The reads mapping and quality control steps were performed through Cellranger (v.7.1.0). The exclusion criteria of cells as below: the number of UMI > 500, percentage of mitochondria gene < 30% and percentage of hemoglobin gene < 5%. After exclusion, we used scDblFinder to detect and remove the cells identified as doublet. The total number of cells included into the final analysis was 106515. A seurat object was generated from the filtered scRNA-seq data using the package Seurat (v.4.1.3). Then we conducted the Single Cell Transform (SCT transform), Principal Component Analysis (PCA), graph-based clustering and Uniform Manifold Approximation and Projection (UMAP) to reduce the dimension information of the scRNA-seq data and clustered the cells into 27 subclusters and annotated the clusters with 11 main cell types with their feature genes. We applied chi-square test to evaluate the difference of cell composition among three CMBRGs and spearman correlation to identify the CMB associated with specific cell clusters. CellChat (v.1.6.1) was employed to describe the ligand-receptor interaction between each cell subclusters. For each CMB, Pearson correlation analysis was performed among all the TCGA pan-GI cohorts to determine the genes significantly associated with each CMB (Statistical significance: *P* value < 0.05). Using the *AddModuleScore* function from package Seurat, we added the score of each CMB based on the average expression level of CMB-related genes to every single cell. UMAP plot was deployed to illustrate the distribution of each CMB score in different cell clusters. The scores of proliferation related genes and G2/M phase related genes were estimated and illustrated using the similar approach.

### Multiplexed immunohistochemical (IHC) staining

5μm Paraffin-embedded sections were deparaffinized with xylene and gradient ethanol solutions, and antigen retrieval was conducted via microwave treatment. Endogenous peroxidase was neutralized with endogenous peroxidase blocking solution (SP KIT-A1 Fuzhou Maixin Biotech. Co., Ltd.) was used to block the binding of irrelevant antibodies. The primary antibodies series included anti-CD4 (Cat No. ab133616, dilution 1:500, Abcam), anti-CD8α (Cat No. 85336, dilution 1:200, Cell Signaling), anti-CD3 (Cat No. ab16669, dilution 1:150, Abcam), anti-CD68 (Cat No. 76437, dilution 1:400, Cell Signaling), anti-CD206 (Cat No. 91992, dilution 1:400, Cell Signaling) and anti-CD80 (Cat No. ab134120, dilution 1: 1000, Abcam) antibodies. Following the application of the Opal polymer HRP anti-rabbit secondary antibodies (Panovue), to detect the antibody staining, and the tissues were incubated with one of the following fluorophores according to the manufacturer’s instructions: PPD 520, PPD 620, PPD 570, PPD 540, PPD 650 and PPD690 (dilution 1:100). The tissue section was then mounted in SlowFade Gold Antifade Reagent with DAPI (Thermo Fisher Scientific, Inc.). Whole slide tissue scanning was performed at 10× magnification using the Vectra Polaris System (Akoya Biosciences), and tumor area according to the H&E staining of the adjacent slide was performed at 20× magnification for subsequent analysis in Inform 2.6.0. The intensity of each fluorescein was deconvoluted from 8 randomly selected fields at 20× magnification. After the acquisition and deconvolution of fluorescence signal, we used spearman correlation to find the CMB related to specific cell cluster.

### Nomogram with calibration curve

To evaluate the capability of CMBRG in predicting the prognosis, nomogram models for each 9 cohorts containing all the independent variables related to the prognosis or recurrence were constructed with package rms (v6.7.1). All these models were validated by calibration curve through 3-fold cross-validation.

### Statistical Analysis

Statistical analysis was performed with the software: R (version 4.2.2). Kaplan-Meier plot with log-rank test was used to evaluate the prognostic value of the CMBRG. Multivariable CoxPH regression model was employed to demonstrate the significant and independent prognostic value of CMBRS by adjusting for other important clinical factors. Pearson or Spearman correlation analysis was used to evaluate the association between CMBRS and other clinical or pathological factors. The receiver operating characteristic curve (ROC) model was employed to describe the performance of prognostic model with or without CMBRS (Statistical significance: *P* value < 0.05).

### Results Study design

To develop tissue agnostic biomarkers from H&E-stained diagnostic slides, we designed a comprehensive and international multicenter study with multi-step sequential evaluation and validation (Figure 1):

a) **Discovery and validation in colon cancer to assess clinical value of CMBs**: We used our well-established CMB-ML to discover CMBs from H&E-stained diagnostic slides, then established a CMB signature and score system using TCGA-COAD cohort. Clinical values of the CMB signature and score system were evaluated with independent validation using the colon cancer patient cohort from Drum Tower Hospital.
b) **Transfer learning to develop tissue-agnostic biomarkers for pan-GI cancer**: We used transfer learning to translate CMBs, the CMB signature and scoring system into other GI cancers and then comprehensively evaluated their clinical values together with independent validation.
c) **Biological understanding of CMBs to provide therapeutic mechanisms and strategies by utility of CMBs:** We discovered biological functions associated with each CMB using bulk RNA and scRNA sequencing with validation by opal multiplex IHC.
d) **Clinical application of CMBs in precancerous lesions to assist assessment, diagnosis and treatment planning**. We deployed clinical application of CMBs, the CMB signature and score system for risk assessment of precancerous lesions and early-stage cancers to help tailor clinical decisions and personalized care.
e) **Integration of multimodal makers to improve predictive power**: We established a strategy for integration of multimodal factors including CMB signature, gene signature and clinical factors to improve risk prediction for precision clinical decisions.

Overall, this study aimed to provide a new AI-based avenue that aids clinicians to make precision clinical decisions by cross-cancer knowledge transfer learning.

### CMB signature discovery and its clinical value in TCGA-COAD

At the initial stage of our research, the CMB-ML pipeline^28^ characterized cellular objects that were represented by 15 morphometric properties from H&E-stained diagnostic slides of 430 patients in TCGA-COAD cohort, and thereafter identified 128 CMBs through unsupervised sparse learning. Subsequently, we profiled each patient as a 128-dimensional feature vector consisting of the relative abundance of 128 CMBs and then evaluated the prognostic value of the 128 CMBs with respect to OS in TCGA-COAD cohort.

OS analysis revealed that 75 of 128 CMBs had a significant prognostic impact in the TCGA-COAD patient cohort (*P* < 0.05). Among them, 13 CMBs (Supplementary Figure S6) demonstrated independent and significant association with OS (Figure 2B), thus defined as a 13-CMB signature for establishing a risk score (CMBRS) for each patient. To assess the prognostic value of such a signature, we constructed the CMBRS and stratified TCGA-COAD patients into three risk groups (High: top third; Intermediate: middle third; and Low: bottom third) based on CMBRS values. We found that these three groups explain significant differences in OS values (*P*< 0.0001, Figure 2B, top panel). Importantly, in the multivariate model to compare CMBRS with clinical factors, CMBRS showed independent of clinical factors (Hazard Ratio (HR)=2.08, *P*= 0.001, Figure 2B, bottom panel). Unsurprisingly, we showed that the integration of CMBRS with clinical factors significantly improved the prognostic power compared to clinical factors or CMBRS only (*P*<0.05; Supplementary Figure S7A, Supplementary Figure S8A, Supplementary Figure S9A). And the calibration curves confirmed the predictive power on prognosis of the nomogram constructed on the CMBRS and clinical factors, enabling convenient and robust clinical application (Supplementary Figure S9B).

**Figure 2.**
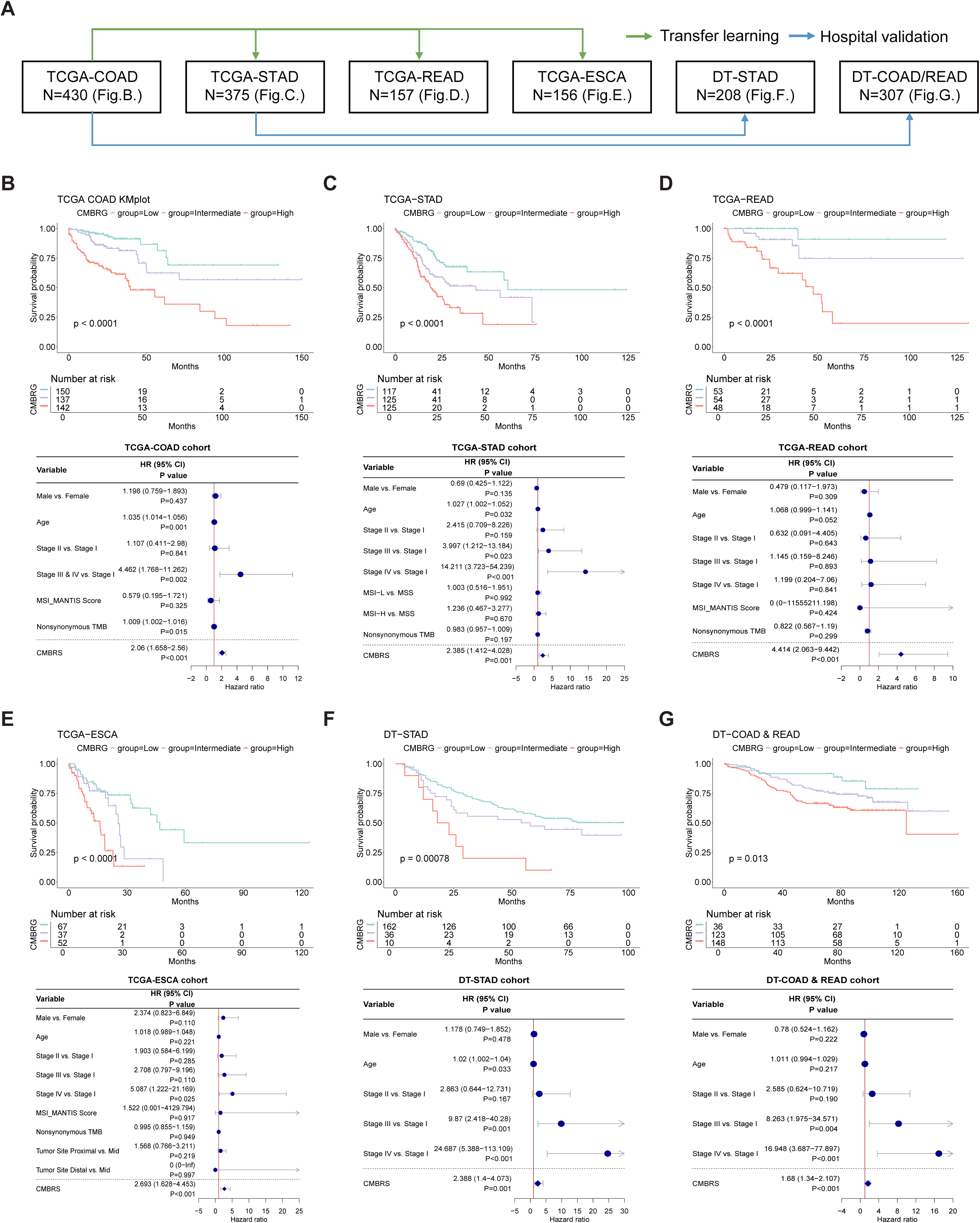
(A) The composition of learning and validation cohort. (B-G) K-Mplots illustrated the predictive effect of CMBRG and forest plots illustrated the independency of CMBRG on 6 cohorts. Statistics performed by log-rank test.

### Evaluation and validation of 13-CMB signature as a tissue**-**agnostic biomarker for different GI cancer types

To test whether the 13-CMB signature developed in TCGA-COAD is a tissue agnostic biomarker for other GI cancer types, we profiled CMBs landscape from WSI of diagnostic slides of TCGA patients with rectal (TCGA-READ), gastric (TCGA-STAD) and esophageal (TCGA-ESCA) cancer and calculated CMBRS based on the pre-built model in TCGA-COAD. Similar to our findings in TCGA-COAD cohort, we found that the CMBRS is a significant prognostic factor independent of clinical factors in these three cancer types (*P* < 0.0001; HR =4.414 in TCGA-READ; HR = 2.385 in TCGA-STAD; and HR = 2.693 in TCGA-ESCA; Figure 2C, D and E). Consistently, the integration of CMBRS with clinical factors significantly improved the prognostic power compared to unimodal systems across cancer types (Supplementary Figure S7B, C and D, Supplementary Figure S8B, C and D, Supplementary Figure S9C, E and G; *P* < 0.05; Justified by AUC and C-Index), and the corresponding multimodal nomogram (Supplementary Figure S9D, F and H) enabled convenient and robust (Supplementary Figure S9D, F and H, justified by calibration curves) clinical application.

To validate the 13-CMB signature as a tissue-agnostic biomarker, we assessed its prognostic value in two independent DT hospital cohorts of 208 patients with stomach adenocarcinomas and 307 patients with CRC (Supplementary Figures S1 and S2). We revealed that the CMBRS was a significant prognostic factor independent of clinical factors in both cohorts (*P* = 0.01, *P* < 0.001; Figures 2F, G, Supplementary Figure S5E, F, Supplementary Figure S6I, L). The generalizability and reproducibility of the CMBRS indicate that the 13-CMB signature is a tissue-agnostic biomarker for pan-GI cancer.

### CMB-related biological function

To decipher the CMB-related biological functions, we identified genes transcriptionally correlated to each CMB in the four TCGA datasets (Supplementary Table S13). We then constructed enrichment networks of Gene Ontology (GO) on biological process (Figure 3A), Kyoto Encyclopedia of Genes and Genomes (KEGG) pathways, Cellular Component and Molecular Function (Supplementary Figure S10) significantly correlated with each of the 13 CMBs in our signature. These analyses revealed that 11 of the 13 CMBs were significantly enriched for five biological functions including: cellular proliferation, immune microenvironment, DNA repair etc. CMB12, CMB67, CMB76, CMB56, CMB97 and CMB120 were associated with the formation of cell junction, cytoskeleton and extracellular matrix. CMB9 and CMB85 were associated with the process of cell replication and DNA repair. CMB29 was associated with the function of innate and adaptive immunity. CMB47 was associated with the regulation of adaptive immunity.

**Figure 3.**
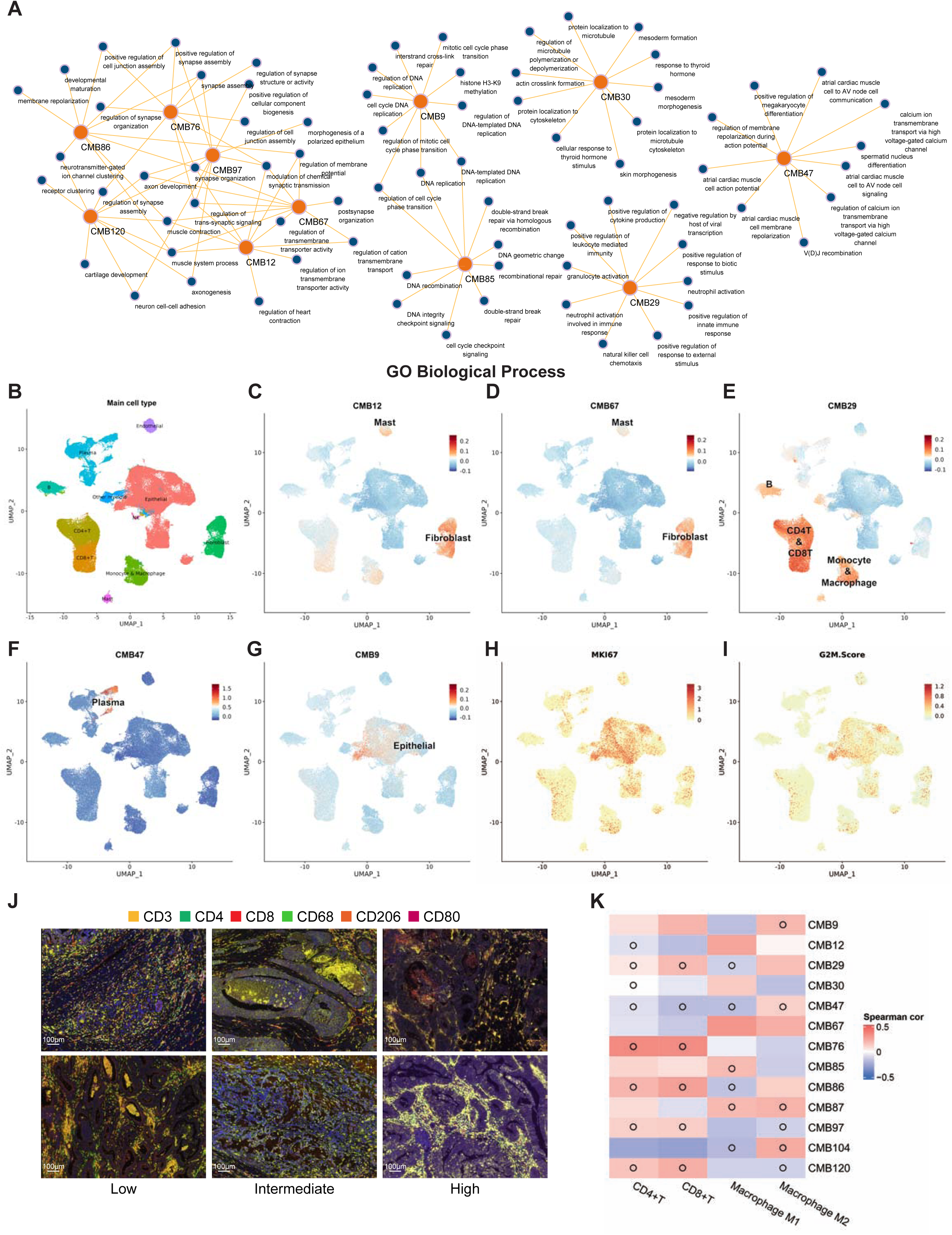
(A) KEGG pathways significantly enriched by the correlated genes of eight CMBs. (B-I) UMAP visualization of single-cell sequencing results, highlighting cell clusters and prognostically representative CMBs marker expression. (J) Multiplex immunofluorescence illustrated the 5 known marker of tumor associated immune cells, CD8+T(CD3+/CD8+), CD4+T(CD3+/CD4+), M1 macrophage (CD68+/CD80+) and M2 macrophage (CD68+/CD206+). (K) Heatmap illustrated the correlation between CMBRS and cellular composition from the multiplex immunofluorescence. Box with a circle indicated that these two methods have consistent results. Statistics performed by spearman’s correlation (* *P* < 0.05)

To further understand the biological relevance of the 13 CMBs, we conducted CMB profiling in combination with single-cell RNA-sequencing of colon cancers prospectively collected from 17 patients in a double-blinded manner and assessed relationships between CMBs and the composition of different cell types. Among these patients, none of them were diagnosed with Hereditary nonpolyposis colorectal cancer or has the family history of colon cancer, and only one patient was identified as Microsatellite Instable (i.e., MLH1 & PMS2 deficiency) using IHC analysis. After quality control procedure, we clustered 106,515 cells into 11 cell groups, and found a significant difference in the composition of different cell types (*P* < 0.001, Figure 3B, Supplementary Figure S11). Next, we estimated the genes transcriptionally correlated to each CMB in these cell types (Figure 3C-G, Supplementary Figure S12). The genes associated with CMB9, CMB85 and CMB104 were predominantly expressed in tumor cells (Figure 3G, Supplementary Figure S12B,F), where CMB9 captured proliferation of tumor cells (Figure 3H-I); the genes associated with CMB12, CMB67, CMB76, CMB86, CMB87, CMB97 and CMB120 were predominantly expressed in fibroblasts (Figure 3C,D, Supplementary Figure S12 A,C,D,E and G); the genes associated with CMB29, CMB86, CMB87, CMB97 and CMB120 were predominantly expressed in T lymphocytes (Figure 3E, Supplementary Figure S12 C,D,E and G); the genes associated with CMB29 and CMB87 were predominantly expressed in monocytes and macrophages (Figure 3E, Supplementary Figure S12D); the genes associated with CMB29, CMB97 and CMB120 were predominantly expressed in B cells (Figure 3E, Supplementary Figure S12E and G); the genes associated with CMB47 were predominantly expressed in plasma cells (Figure 3F); and the genes associated with CMB12, CMB67, CMB86, CMB87, CMB 97 and CMB120 were predominantly expressed in mast cells (Figure 3C and D, Supplementary Figure S12C, D, E and G). Moreover, using CellChat, we revealed the CMB-specific association with cell type interactions (Supplementary Figure S13). For example, CMB29 is positively associated with the interactions among different type immune cells. Furthermore, we verified the association between CMBs and immune cell types by multiplexed IHC staining on FFPE samples from DT-COAD-scRNASeq cohort (Figure 3J-K). Notably, the biological interpretation of CMBs by scRNA-seq and multiplex IHC staining are highly consistent with pathologic interpretation by pathologists (Figure 4), including the CMBs that captured distinct cell types such as tumor cells (e.g., CMB12, CMB85, CMB104), fibroblasts (e.g., CMB67, CMB76, CMB86, CMB87, CMB9), immune cells (e.g., CMB29, CMB47), and immune- and stroma-rich microenvironments (e.g., CMB97, CMB76, CMB120).

**Figure 4.**
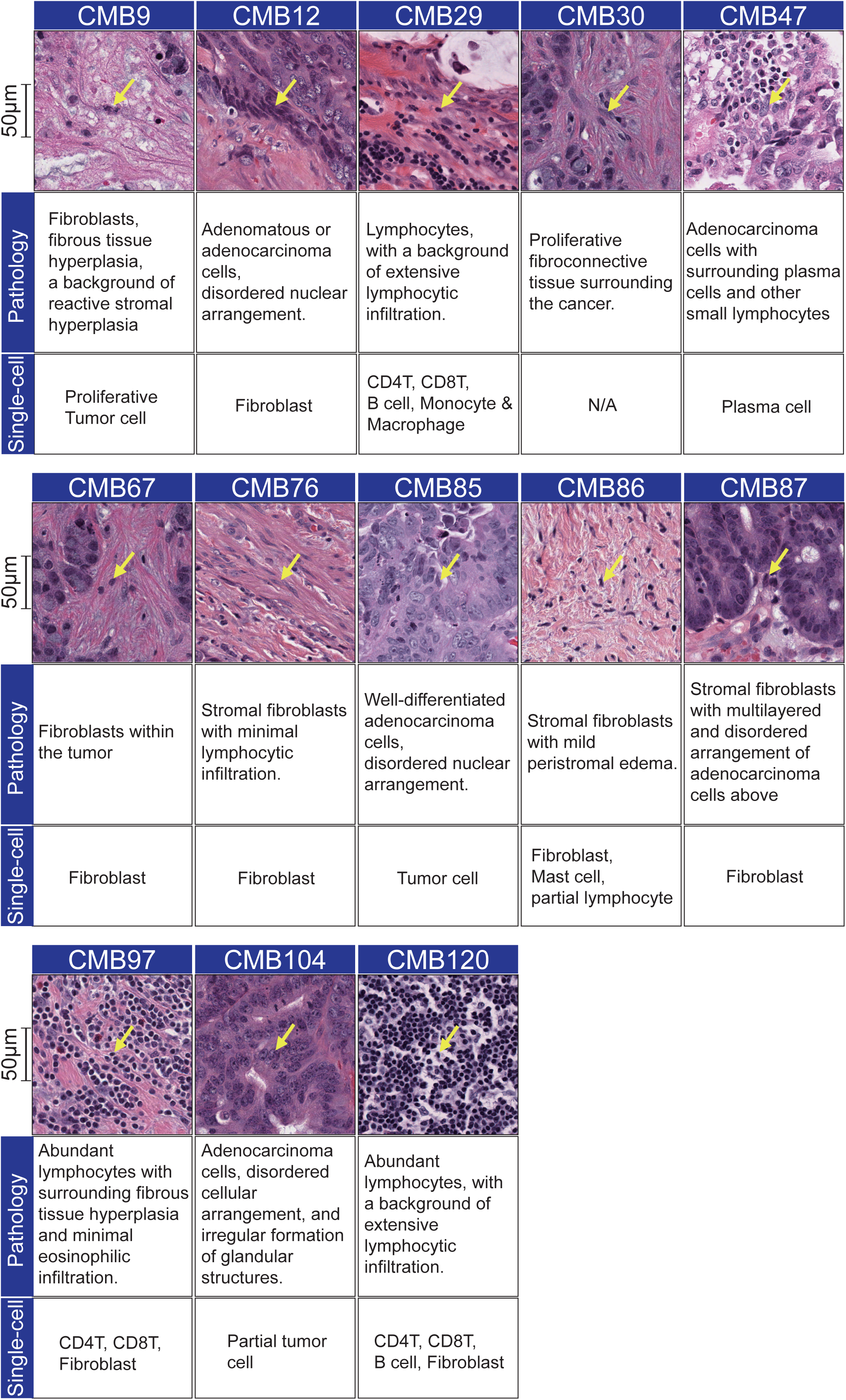
Representative pathological images of 13 prognostically significant CMBs, along with their pathological and single-cell sequencing interpretations (highlighted with yellow arrows).

Taking all the findings from bulk RNAseq, scRNAseq, Opal staining, and pathologic interpretation, we conclude that CMBs pose consistent biological and functional interpretability.

### Clinical application of 13-CMB signature to risk stratification of precancerous lesions and early-stage cancer

Risk stratification of early-stage cancer is essential for guiding clinical decisions. Thus, we conducted evaluation of the 13-CMB signature for risk stratification in two common clinical scenarios: colon adenoma and early esophageal neoplastic lesion (Figure 5A, Supplementary Figures S4 and S5). The KM-plot with log-rank test exhibited that the CMBRG was significantly associated with recurrence-free survival, where the median recurrence-free survival in low CMBRG is 42.5 months while it is 23.7 months in high CMBRG (Figure 5B). Importantly, CMBRS is an independent factor for risk stratification by adjusting for the clinical factors, including age, gender, location, number of adenomas, etc. (Figure 5C). In addition, CMBRS significantly outperforms clinical recurrent factors in risk prediction and further enhances the predictive power through the integration of multimodal factors (e.g., clinical recurrent factors and CMBRS) (Figure 5D). A nomogram (Figure 5E top panel), integrating CMBRG and other recurrent factors, including gender, age, advanced adenoma, lesion site and total number of adenomas, enables accurate (Figure 5E bottom panel, justified by calibration curve) and convenient clinical deployment. All these findings were further validated in an independent DT-CAP-Validation cohort (Figure 5F-I).

**Figure 5.**
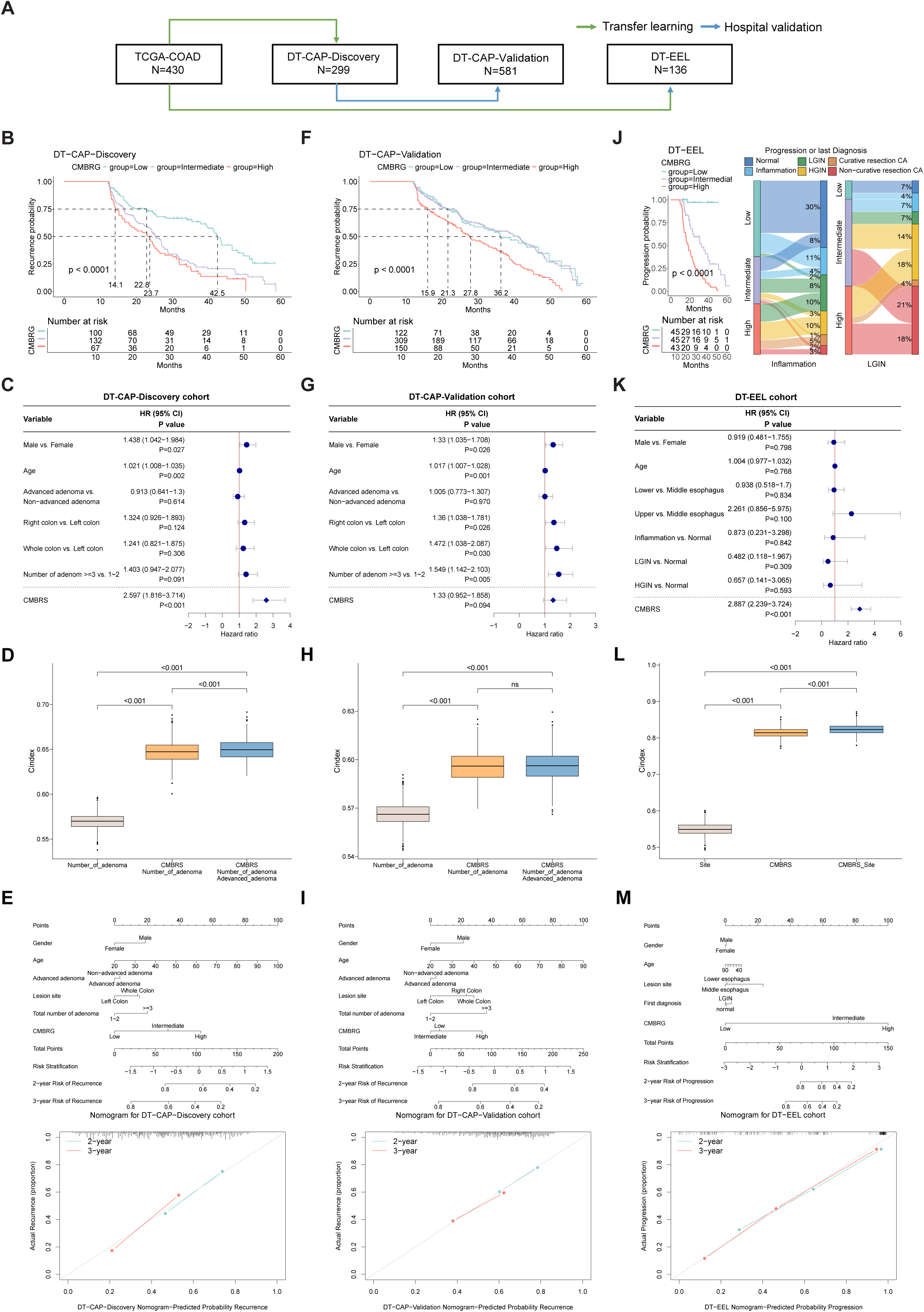
(A) The composition of learning and validation cohort. (B, F, J) K-Mplots illustrated the predictive effect of CMBRG and (C, G, K) forest plots illustrated the result of the multivariate cox regression on CMBRG and prognostic factors. Statistics performed by log-rank test. (J) The Sankey Plot visualization of the different progression of the inflammation and LGIN patients from DT-EEL cohort, among three CMBRGs. Boxplots illustrated the C-index of different combinations of prognostic factors after 1000-step bootstrap and nomograms with calibration curve predicted the 3-year and 5-year survival probability among (D, E) DT-CAP-Discovery, (H, I) DT-CAP-Validation, (L, M) DT-EEL cohorts. Statistics performed by t-test.

Similarly, CMBRG could also stratify patients with early esophageal neoplastic lesion into risk groups that predict diseases progression independent of clinical factors (Figure 5J-M). Specifically, over 95% of the patients with esophageal inflammation in low CMBRG never progressed with nearly 70% recover rate (e.g., from inflammation to normal) (Figure 5J, middle panel); while 100% of the patients with esophageal inflammation in high CMBRG have disease progression (Figure 5J). Same observations were found in patients with esophageal LGIN. Specifically, none of the patients with LGIN in low CMBRG progressed, while 100% of the patients with LGIN in high CMBRG have disease progression (Figure 5J, right panel).

Taking all together, these findings suggest clinical utility of the 13-CMB signature for personalized management for patients with early-stage cancerous lesions.

### Integration of CMB and gene signature to improve predictive accuracy

Lastly, we investigated whether integration of multimodal biomarker signatures, i.e., CMB and gene signatures, could improve the predictive accuracy of clinical outcomes. In our previous studies, we developed a 15-gene expression signature for colon adenocarcinoma^29^ and a 53-gene signature for gastric adenocarcinoma^30^. A multivariate Cox regression analysis revealed that CMB and gene signatures were independent prognostic factors even after adjusting for clinical factors in both TCGA and independent hospital cohorts (Figure 6A-D). The C-Index showed that integration of clinical stage, CMB signature and gene signature significantly improved prognostic prediction (Figure 6E-H). To further assess the clinical value of the integration of CMB and gene signatures, we established a nomogram model, a valuable clinical tool for prognosis prediction, to predict the 3- and 5-year OS probability of patients. In DT-STAD cohort model, we included gender, age, stage, 53-gene prognostic score and CMBRG. Similarly, we included gender, age, stage, 15-gene prognostic score and CMBRG in the nomogram model in DT−COAD & READ cohort (Figure 6I-L). The calibration curves showed that the predicted value of the 3- and 5-year OS rate by the nomogram was in good agreement with the actual observed value of the 3- and 5-year OS rate (Supplementary Figure S14).

**Figure 6.**
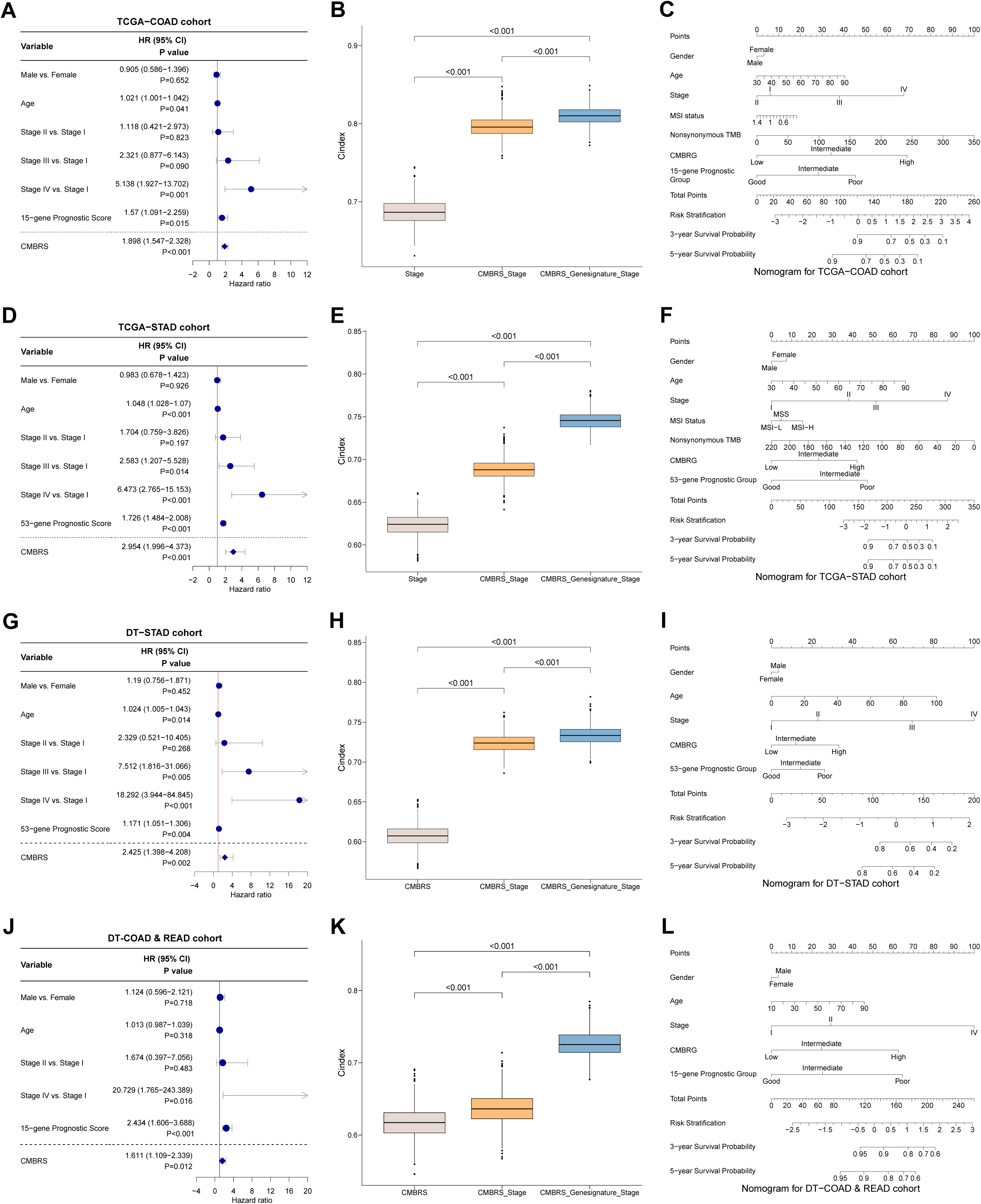
Forest plots illustrated the result of the multivariate cox regression on CMBRG and prognostic factors, boxplots illustrated the C-index of different combinations of prognostic factors after 1000-step bootstrap and nomograms predicted the 3-year and 5-year survival probability, among (A, B, C) TCGA-COAD, (D, E, F) TCGA-STAD, (G, H, I) DT-STAD, (J, K, L) DT-COAD & READ cohorts. Statistics performed by t-test.

Collectively, we conclude that integration of multimodal biomarkers such as CMB and gene expression signatures identified in our own studies can serve as promising tools and future directions to enhance prognostic predictions.

## Discussion

Tissue-agnostic classification of cancers according to their molecular and/or pathologic characteristics not only helps expedite the access of millions of people to effective treatments, but also facilitates a deeper biological understanding of cancer biology with the potential to restructure oncology. As an essential component of tissue-agnostic precision oncology, tissue-agnostic biomarkers, yet underdeveloped, are transformative tools that have the potential to revolutionize cancer care. In this proof-of-concept study, we utilized our previously established AI-pipeline ^17–19^ to discover tissue-agnostic biomarkers characterizing cellular morphometric heterogeneity and commonality from the WSIs. Our comprehensive findings on different clinical risk assessments strongly demonstrated the clinical value of this AI-empowered 13-CMB signature as a tissue agnostic biomarker for management of pan-GI pre-cancer and cancer patients. Also, we presented evidence that CMBs are synergistic to the existing molecular biomarkers identified by our researchers, i.e., the multi-gene prognostic signatures in colon cancer ^29^ and gastric cancer ^30^. Moreover, we provided biological insights into CMBs using bulk RNA-seq and validated them using both scRNA-seq and Opal Multiplex IHC, which leads to our understanding of therapeutic mechanisms and strategies by utility of CMBs. Our study lays the foundation for future implementation of AI-powered CMBs in tissue-agnostic precision oncology research and clinical practice and provides a more cost-effective and accessible option for cancer patients and clinicians. Development of biomarkers from H&E-stained tissue sections benefits greatly from recent advancements in AI and has rapidly expanded. From a technical point of view, these works have largely focused on end-to-end prediction of clinical outcomes and/or molecular characteristics, leaving the biomedical understanding and justification of these AI-derived biomarkers, especially at both single cell transcriptome and protein levels, insufficient addressed. In contrast, in this study, we also provided biological interpretability of CMBs, which reveals that CMBs are in line with previous molecular knowledge of cancer prognosis and explain the robustness and effectiveness of AI-discovered CMBs in prognosis prediction of cancer patients. Moreover, consistent with previous reports, we found that CMB signature is an independent prognostic factor compared with gene signatures and clinical factors, suggesting that CMBs can provide additional clinical value complementary to that provided by the current clinical and molecular techniques. This also led us to test the possibility of improving prognostic predictions by integration of multi- modal biomarkers such as CMB and multi-gene signatures. Indeed, our results demonstrated the superiority of multi-modal biomarker integration for prognostic prediction as compared with unimodal biomarkers alone. In future studies, we plan to link CMBs to treatment responses of different therapeutic agents and evaluate/establish the role of CMBs in clinical decision-making for an effective cancer treatment.

Biotechnological advancement enables early detection of cancer, significantly improves the chances of successful treatment, and reduces mortality. However, the guideline for clinical management of early-stage cancer has not yet been fully established which leads to overtreatment of cancer patients ^31^. For example, in current clinical practice, patients with adenomas are all suggested to undergo excessive colonoscopy surveillance post-polypectomy, among whom only 6% develop CRC, which imposes big challenges in clinical management ^32–34^. Therefore, precision risk assessment of early-stage cancer is an urgent need. To this end, we developed the CMBRS to assess the recurrence risk of colorectal adenoma, where the median recurrence-free survival in low CMBRG (around 40 months) is significantly longer than that in high CMBRG (around 25 months), suggesting the possibility of precision clinical management of patients per CMBRG. Specifically, by refining the surveillance schedule based on individual risk assessments, the scoring system can help prevent unnecessary, overly frequent follow-ups for patients with low-risk adenomas, thereby optimizing healthcare resources and minimizing patient burden. Importantly, this scoring system enables robust, cost-effective and rapid clinical implementation worldwide.

The AI-powered CMB predictive model in this study also offers a new tool for early detection of esophageal squamous cell carcinoma (ESCC) and therefore personalized management of early esophageal lesions (EELs). Unlike traditional diagnostic methods that primarily assess the current state of the disease ^35–37^, our model provides valuable foresight into the potential progression of EEL, including whether the lesion is more likely to evolve into curative or non-curative cancer. By assessing the risk of progression from initial diagnosis, the CMBRS model revealed that in the low CMBRG, over 95% of patients never progressed, among whom over 60% of patients recovered from diseases. In contrast, 100% of the patients in the high CMBRG had disease progression. This allows more targeted and proactive management. For patients in low CMBRG, our findings suggest conservative monitoring without the need for immediate invasive procedures, but patients in higher CMBRG require more intensive monitoring and early intervention even before the lesion progresses to invasive carcinoma, such as endoscopic submucosal dissection (ESD). Overall, the AI-powered CMBRS model provides a significant step forward in the personalized care of patients with EEL, offering a tool to improve survival rates while minimizing overtreatment.

Our study brings valuable insights and significant contributions to the AI-based biomarker field, but we are also aware of some limitations of this study. For example, retrospective cohort studies were used for training and validation, so the future studies should pursue prospective multicenter clinical evaluation of the 13-CMB signature. Secondly, we primarily focused on prognostic evaluation of the CMBs, although such information is very useful for clinical management of cancer patients, we will also evaluate the predictive power of CMBRS model on therapeutic/interventional outcomes in future studies.

In summary, we for the first time developed, extensively validated and biologically interpreted an AI-powered tissue-agnostic CMB signature from WSIs that highlights the untapped potential of its clinical application in management of patients with GI precancerous lesions or cancers. Furthermore, we showed that integrating multimodal biomarkers, e.g., CMB signature and multi-gene signature, facilitates better stratification of cancer patients for precision oncology, suggesting the independent clinical value of both molecular and pathological profiling. Finally, we demonstrated that CMBs can capture commonalities across different types of precancerous lesions and cancers in GI tract, which potentially provides a new avenue for tissue-agnostic precision care.

## Data Availability

Whole slide images and clinical data of the TCGA cohorts were downloaded from the TCGA GDC portal (https://portal.gdc.cancer.gov/). Processed data related to TCGA and Drum Tower cohorts have been provided with the manuscript. Raw data from the Nanjing Drum Tower Hospital is not currently permitted in public repositories because ethical and legal implications are still being discussed at an institutional level.

## Author Contributions

JF, LW, JHM and HC conceptualized and designed the study. AWM led the model construction and formal analysis from public cohorts. PW and JCF led the independent hospital validation, processed the raw data and performed formal analysis in hospital validation cohorts. QS provided pathological interpretation. PW, CFJ, AWM, QS, HZ, JI, SC, AMS, DWT, AB, BH, JF, JHM, LW, and HC were involved in the critical review of the data and/or interpretation of results. PW, CFJ, AWM, JF, JHM, LW and HC drafted the manuscript. All authors edited, reviewed, revised, and approved the manuscript text.

## Acknowledgements

the authors would like to thank Lei Xu, Binbin Yuan, Lingyan Chen, Yingjia Zhuang, and Jingjing Wei for their contributions to cohort data collection, follow-up, pathology slide retrieval and scanning, and surgical specimen acquisition.

## Abbreviations

H&E: hematoxylin and eosin
AI: artificial intelligence
GI: gastrointestinal
CMB: cellular morphometrics biomarker
WSI: whole-slide images
CMBRS: CMB risk score
CMBRG: CMB risk group
OS: overall survival
scRNA-seq: single cell RNA sequencing
IHC: immunohistochemistry
TCGA-COAD: The Cancer Genome Atlas - Colon Adenocarcinoma
TCGA-STAD: The Cancer Genome Atlas - Stomach Adenocarcinom
TCGA-READ: The Cancer Genome Atlas - Rectum Adenocarcinoma
TCGA-ESCA: The Cancer Genome Atlas - Esophageal Carcinoma
LGIN: Low-Grade Intraepithelial Neoplasia
HGIN: High-Grade Intraepithelial Neoplasia
CAP: Colon Adenomatous Polyps
EEL: Early Esophageal Lesion
CRC: Colorectal Cancer
DT: Drum Tower Hospital

